# Do Public Health Efforts Matter? Explaining Cross-Country Heterogeneity in Excess Death During the COVID-19 Pandemic

**DOI:** 10.1101/2022.11.21.22282563

**Authors:** Min Woo Sun, David Troxell, Robert Tibshirani

## Abstract

The COVID-19 pandemic has taken a devastating toll around the world. Since January 2020, the World Health Organization estimates 14.9 million excess deaths have occurred globally. Despite this grim number quantifying the deadly impact, the underlying factors contributing to COVID-19 deaths at the population level remain unclear. Prior studies indicate that demographic factors like proportion of population older than 65 and population health explain the cross-country difference in COVID-19 deaths. However, there has not been a holistic analysis including variables describing government policies and COVID-19 vaccination rate. Furthermore, prior studies focus on COVID-19 death rather than excess death to assess the impact of the pandemic. Through a robust statistical modeling framework, we analyze 80 countries and show that actionable public health efforts beyond just the factors intrinsic to each country are necessary to explain the cross-country heterogeneity in excess death.

## 1 Introduction

The World Health Organization (WHO) estimates that the COVID-19 pandemic has led to 14.9 million excess deaths worldwide in 2020 and 2021 [1]. Excess death is defined as the difference between actual reported death counts and what was expected under “normal” conditions based on data from earlier years [2]. Excess death has shown to be a more accurate measure assessing the true impact and death toll of the COVID-19 pandemic [3–5]. For instance, in the United States, a death is registered as death by COVID-19 only if the patient had a positive test result. However, early in the pandemic when resources were scarce and hospitals were overwhelmed, it is likely that COVID-19 tests were unavailable for these patients.

Although the world collectively experienced a massive number of these excess deaths during the COVID-19 pandemic, the impact across different countries varies widely. While some countries suffered around 450 excess deaths per 100, 000 people, other countries saw nearly 0 excess deaths per 100, 000 [1]. What factors drive this change? Were certain countries simply better suited to handle the pandemic due to robust healthcare infrastructure, a healthier population, and a younger population? Or can these pre-existing traits not explain such variations alone, implying that some COVID-19 related public health efforts lessened the impact of the pandemic? In this paper, we split the set of all covariates into two distinct subsets: “intrinsic” features that countries inherited before COVID-19 was declared a pandemic in March 2020, and “actionable” features that can potentially be modified within the timeframe of the pandemic. By explicitly investigating governmental policies and public health efforts in conjunction with ingrained economic and health factors through this lens, we can better understand and explain the drivers of excess deaths and the large variations of fatalities between countries.

Much of the past work regarding COVID-19 death at the country-level focuses on the impact of the pandemic on groups in different income levels (such as [6], [7]). Another major area of study is the thorough analysis of COVID-19 death drivers in specific, single countries (such as [8], [9]). In regard to studying the variation in COVID-19 deaths across countries, articles like [10] reveal that being strict in regards to certain policies - such as testing and contact tracing policies - are highly associated with countries that initially mitigated the spread and effect of the disease. However, this study and many others do not place a quantitative comparison among these factors, and no statistical model is created to estimate these features’ impacts. Other studies purely investigate intrinsic population risk factors. For example, the findings from [11] state that Alzheimer’s disease and some lung-related illnesses are associated with higher case mortality rates of COVID-19. The study performed in [12] incorporates both population risk factors such as lung-related illness prevalence and governmental policies like COVID-19 testing strategies. However, these models predict case fatality rate (CFR) rather than excess deaths. Additionally, like many of the other aforementioned studies, the analysis was performed in 2020 or early 2021 making it impossible to estimate and incorporate the effects of the COVID-19 vaccine.

In our paper, we incorporate and add to many of the characteristics of the studies previously mentioned. Our primary contributions to the study of COVID-19 death variations are as follows:

1. We build a gradient boosting model that allows for the quantification of how much various factors drive excess death. Additionally, we use these models to discuss which combinations of factors have especially deleterious effects on low-GDP countries and relatively unhealthy countries.
2. We view the analysis through the lens of intrinsic features and actionable features, and we specifically demonstrate the prediction power gained from considering actionable public health efforts. We formalize this notion via a bootstrapped hypothesis testing procedure. Also, we fit two models – one with only intrinsic features and one including both intrinsic and actionable features. We use these two models to construct confidence intervals regarding the change in excess death estimates for specific countries after adding actionable variables.
3. We make steps to ensure that our analysis is robust. For example, we use excess death as our target variable which provides a more encompassing view of the pandemic’s mortality burden on a given country. We also incorporate many countries in our study rather than focusing on a particular country or region. Lastly, we incorporate newer, measurable effects such as vaccination rate.

In summary, we uncover a number of important characteristics that are predictive when estimating excess death across nations. Vaccination rate, obesity percentage, and age distribution are three such features. Additionally, intrinsic country characteristics alone cannot explain the large variation in excess death we see across countries. Incorporating actionable features such as vaccination rates and the trust citizens have in their national government’s COVID-19 advice can significantly improve excess death predictions. Across our modeling architecture, we see that relatively obese countries experience disproportionately higher excess death estimates when failing to obtain a higher vaccination rate as compared to low-obese countries. Similarly, low-GDP countries incur elevated excess death estimates when not trusting their government’s COVID-19 advice compared to higher-GDP countries.

## 2 Detailed Results

In this section we report the insights from our statistical analyses. We demonstrate that actionable factors — particularly COVID-19 vaccination rates and trust in official COVID-19 advice from governmental entities — are important in explaining the cross-country heterogeneity of excess death.

### 2.1 Key Drivers of Excess Death

We show in section 4 that our final model successfully captures some of the intrinsic structure and signal in the dataset. The two questions of interest when dissecting how this model makes predictions are “which features do the final model deem as useful?” and “how exactly do these useful features contribute to the model?”. Feature importance methods implemented via the scheme detailed in section 4.2.4 are one way to answer the first question. Figure 1 reveals the six features with the highest relative importance in the final model. Two of the six features are “actionable” features. While explored further in section 2.2, this suggests that the variation in excess death across countries cannot fully be explained via intrinsic variables alone and that some improvement in predictive performance is gained from these actionable variables. The percent of citizens with at least one dose of a COVID-19 vaccine in the study’s time frame and whether citizens base their pandemic-related decisions on advice from their government are factors that could be altered via public health efforts.

**Fig. 1.**
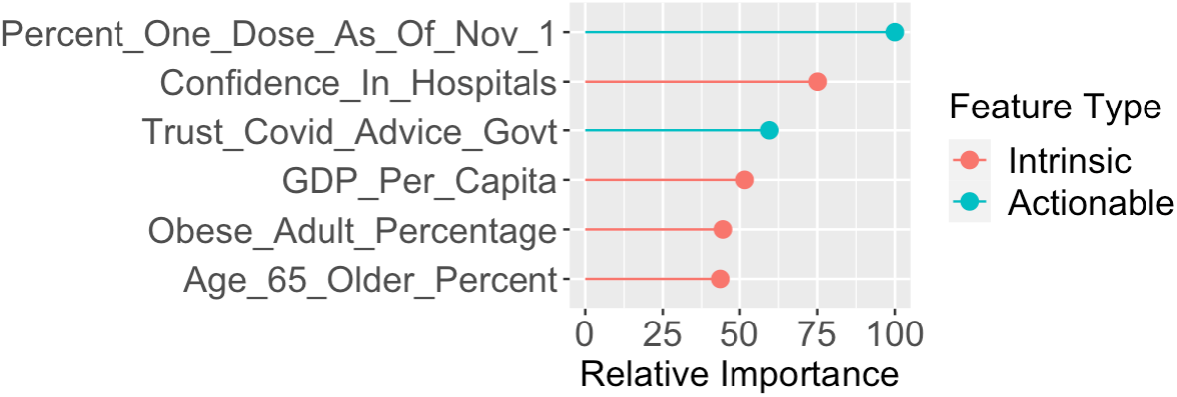
Six features in the final gradient boosting model with the highest relative feature importance.

How do these important features specifically contribute to the model? Figure 2 depicts the partial dependency graphs for the six features with highest relative importance in the gradient boosting model. We see that after accounting for the average effects of all other variables, the estimate of excess death in the model decreases as certain variables increase. These variables include the percent of citizens with at least one COVID-19 vaccine dose, the percent of citizens basing their pandemic-related decisions on official advice from their national government, the percent of citizens who have confidence in their nation’s hospitals, and the GDP per capita. Conversely, the model’s estimate of excess death increases as other features increase. These features include the obesity percentage in the country and the percent of people aged 65 and over. Additionally, we note that there appears to be certain thresholds or “tipping points” in which the model’s estimates begin to widely change. About 60% of citizens receiving at least one COVID-19 vaccine dose and 65% of citizens basing pandemic-related behavior decisions on government’s official advice are tipping points associated with lower excess death estimates. Through these partial dependency plots, we can also look at countries that are similar in numerous aspects yet exhibit a large difference between their excess death. For example, we study the differences in excess death in Canada (54 excess deaths per 100,000 people) and the United States (265 excess deaths per 100,000 people). We see in Figure 3 some explanation as to why the boosting procedure predicts lower excess death estimates for Canada.

**Fig. 2.**
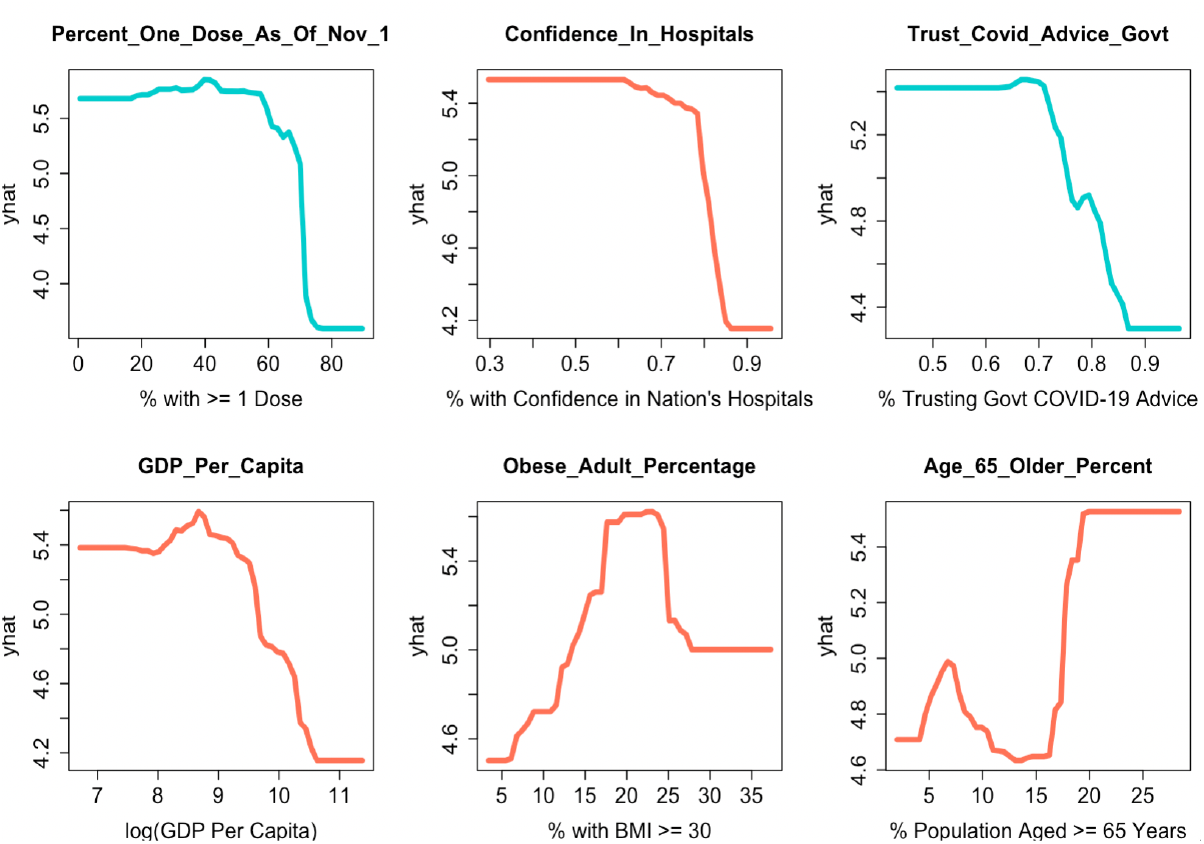
Partial dependency graphs for the six features with the highest relative importance in the final gradient boosting model. The y-axis represents the contribution to the estimation process of excess death (*ŷ*) standardized via a cube-root transformation.

**Fig. 3.**
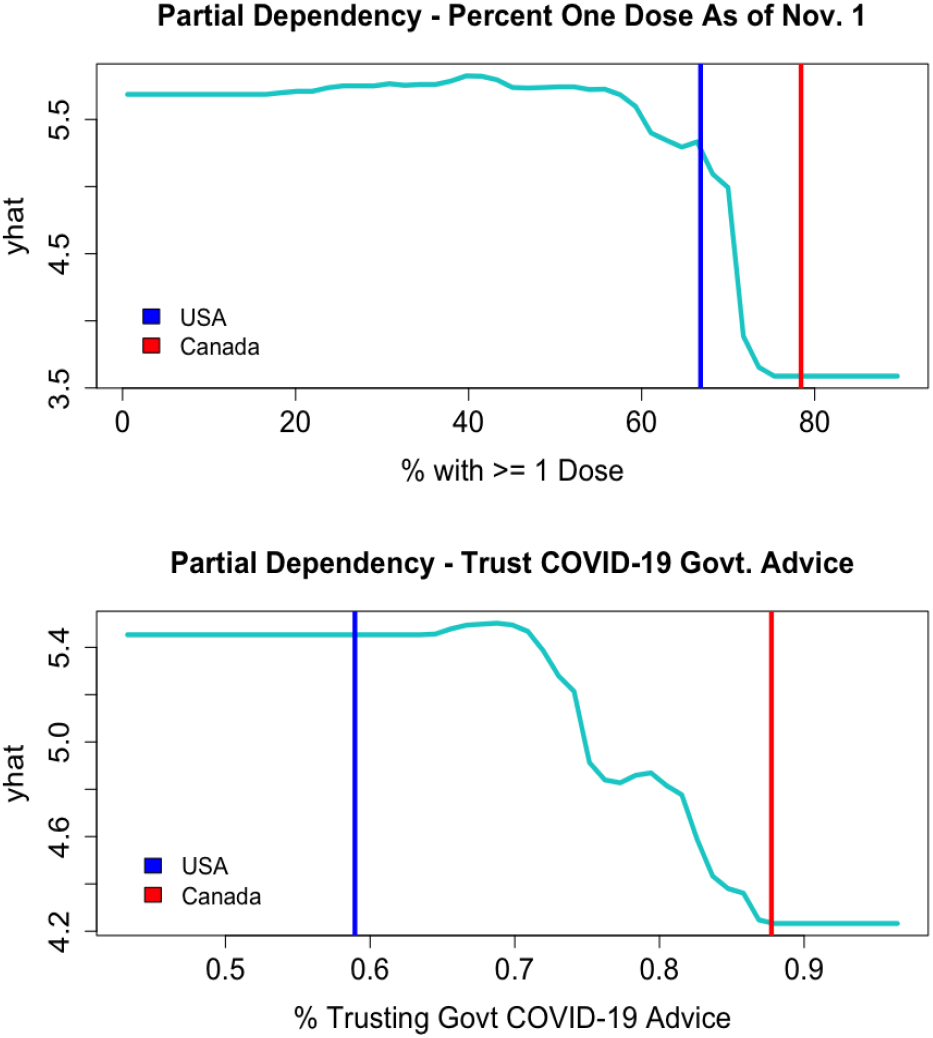
Partial dependency plots for two actionable features. The vertical lines represent the observed values for the United States in blue and Canada in red. The y-axis represents contribution to the (cube-root) estimate of excess death.

After knowing which features are most important to the model and how they contribute to the estimation process, the next question to investigate is “how do variables in the model interact with one another?” Partial dependency plots can provide some insights to this question. For our final gradient boosting model, we see in Figure 4 an interaction between GDP per capita and how much the citizens of a country trust and base pandemic-related decisions on their government’s advice regarding COVID-19. Effectively, we see that the “penalty” - or increase in excess death estimate - for not trusting governmental COVID-19 advice is 43.7% greater for lower GDP countries than higher GDP countries. This is according to this class of models that successfully captures some inherent structure in the dataset as seen in Table 1 and Figure 9. To see this relationship, one can imagine slicing the figure along a (log) GDP per capita of 10.5. Along this curvature, we note the increase in partial dependence as we move from around 85% government trust to around 60%. We see a similar increase exist when slicing the figure at a (log) GDP per capita of 7.5. If no interaction were present, these increases would be nearly equivalent. However, we see the penalty for moving from 85% trust (or higher) to 60% trust (or lower) is greater for countries with 3500 (USD) GDP per capita or less compared to countries with 36000 (USD) per capita or greater. This implies that following official, governmental advice regarding COVID-19 becomes particularly more imperative for low-GDP countries.

**Table 1.** Results of the repeated cross validation scheme for 3 model classes. Hyperparameter grids had similar number of combinations and granularity level across classes.

|  | Gradient Boosting | Random Forest | LASSO |
| --- | --- | --- | --- |
| Best Hyperparameters | Number of Trees = 1600<br>Shrinkage = .007<br>Interaction Depth = 2<br>Min Obs per Node = 10 | Variables Sampled<br>per Split = 3 | $\lambda = .01$ |
| CV RMSE for Best<br>Hyperparameter Combination | 145.2 | 154.3 | 179.6 |

**Fig. 4.**
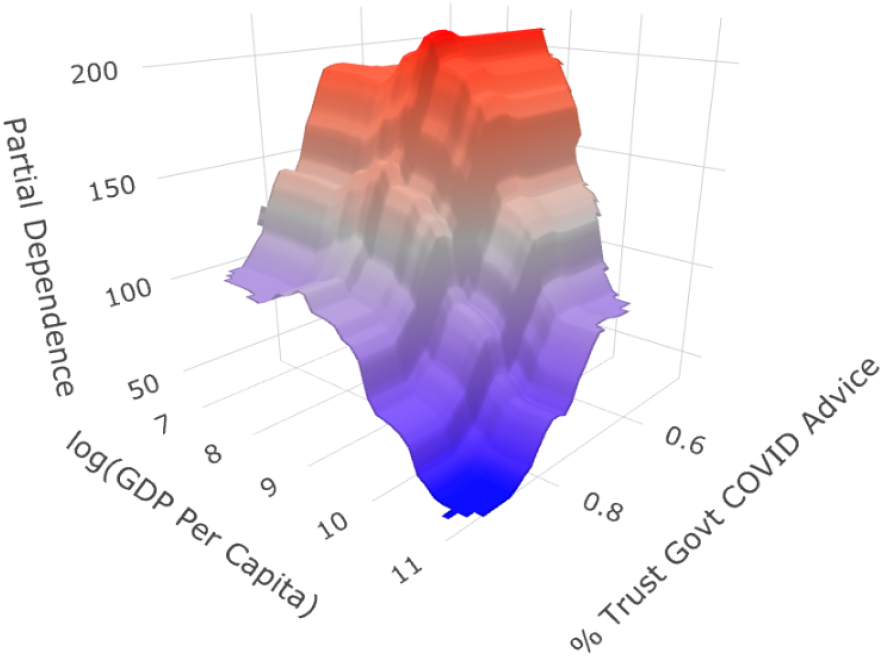
Partial dependency graph for GDP per capita and percent of citizens who base pandemic-related behavior decisions on official advice from their national government.

Next, we can analyze and quantify the interaction between vaccination rate and obesity percentage in the same way. Figure 5 depicts this relationship. Following the same logic as the previous example, we see that countries in roughly the highest obesity quartile experience about a 11.2% greater penalty than countries in the lowest quartile when moving from “very vaccinated” (about 72% vaccination rate or better, which is the top third) to “relatively unvaccinated” (about 50% vaccination rate or worse, which is around the bottom half in our study). Therefore, we see via this quantification how much more imperative it becomes for more obese countries to distribute vaccines to prevent excess deaths compared to less obese countries.

**Fig. 5.**
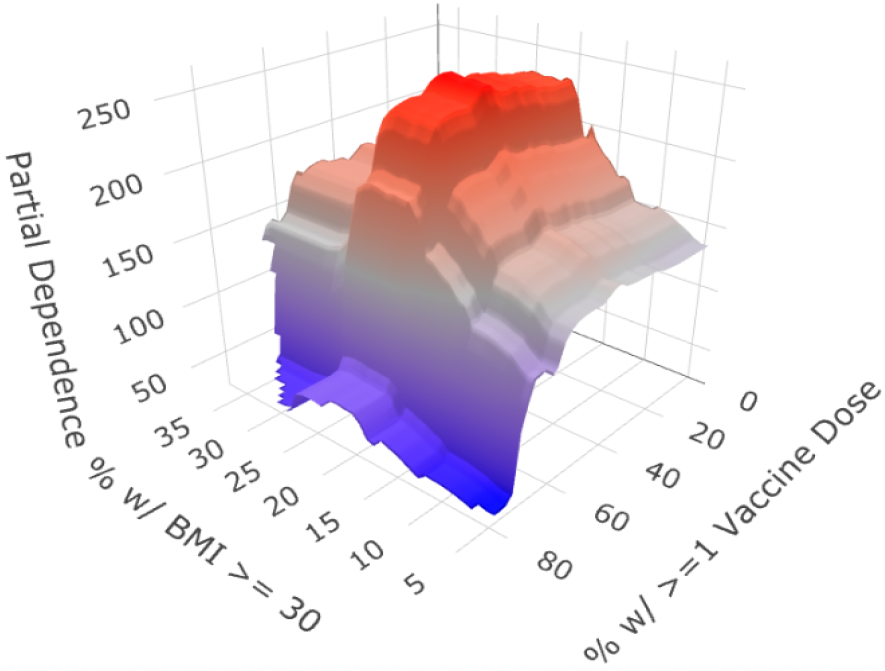
Partial dependency graph for percent of people with at least one dose of COVID-19 vaccine and and percent of citizens with BMI of at least 30.

These partial dependency plots provide a view into the inner-workings of our gradient boosting model. Not only do we see the degree to which certain actionable features decrease excess death estimates, but we also identified particularly harmful combinations of actionable and intrinsic country characteristics. This provides some insights as to which policies and metrics become increasingly important for countries that meet certain criteria.

### 2.2 Increase in Predictive Performance from Actionable Features

Since the final model suggests actionable features are important in predicting country-level excess death, we attempt to formalize this importance via the hypothesis test detailed in section 4. The results from the test indicate that the increase in predictive performance from the set of actionable features is statistically significant (p-value = 0.0001 and observed RMSE difference = 11.7 excess deaths per 100,000 people).

As detailed in section 4, the gradient boosting model gives a statistically significant decrease in RMSE when including actionable features. In other words, the model successfully captures some signal with these features and how they contribute to excess mortality. We also formalized this notion via the aforementioned bootstrapped hypothesis test. A naturally arising question is then “which specific countries see increases in excess death estimates after adding actionable features”? To answer this question, we create a measure called the “delta value”. This value measures the difference in predicted excess mortality between the model with the actionable features and the model without the actionable features, further described in 4. Through these delta values, we can get a sense of which countries “underachieved” with their actionable features during the pandemic. Here, we construct the confidence intervals based on 100 bootstrap iterations. Figure 6 displays the effect of adding actionable features in the model for the 51 countries in which the direction predicted excess death improved. In other words, if *Ŷ*_*I*_ *< Y* then *Ŷ*_*I*_ *< Ŷ*_*IA*_. Or, if *Ŷ*_*I*_ *> Y* then *Ŷ*_*I*_ *> Ŷ*_*IA*_, where *Y* is the observed excess death, *Ŷ*_*I*_ is the predicted excess death from intrinsic-only model, and *Ŷ*_*IA*_ is the predicted excess death from the full model including actionable features. We focus on the top two actionable features identified in section 2.1. We see that countries to the right of the vertical zero line - i.e. countries that have increased predicted excess death after including actionable features - tend to have lower vaccination rates and less trust in COVID-19 advice from governments. The opposite effect holds for countries to the left of the vertical zero line. These countries had lower predicted excess death after including the actionable features and had higher vaccination rate and trust in COVID-19 advice from governments. This further demonstrates the importance of considering the actionable features in understanding the heterogeneity in excess death at the country level.

**Fig. 6.**
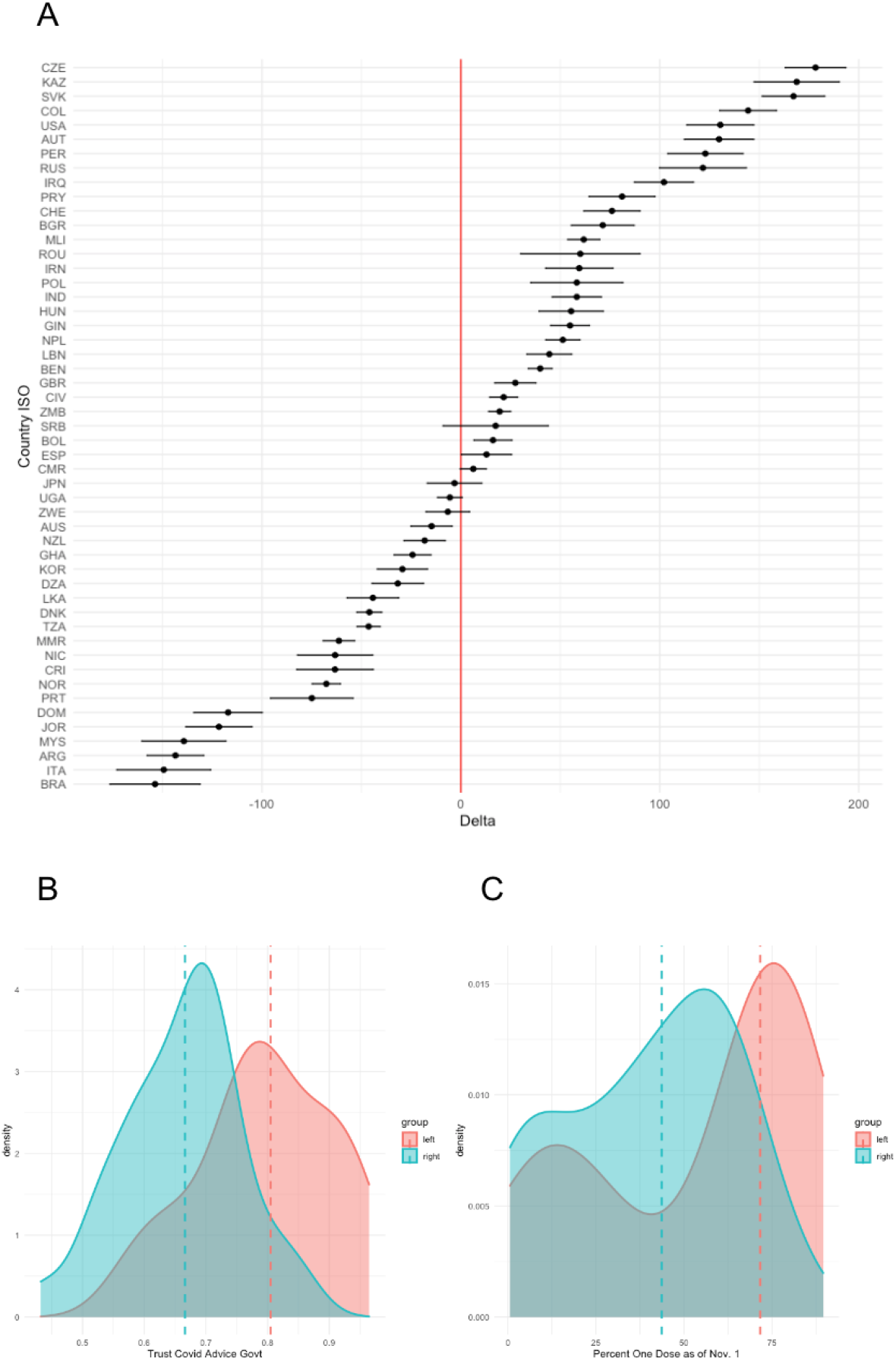
(A) We show the 95% bootstrap confidence intervals of the “delta value”, which measures the difference in excess death prediction between the model fit on both intrinsic and actionable features, and the model fit only on intrinsic features. (B) and (C) are density plots of “trust in COVID-19 advice form governments” and “percent one dose vaccination as of Nov. 1” respectively, colored by whether a country’s delta value lies to the left or to the right of the vertical zero line. The dashed lines represent the median values. Countries with delta to the left of the vertical zero line (i.e. countries with lower predicted excess death) tend to have higher vaccination rate and trust in government COVID-19 advice.

## 3 Discussion

In December 2019, the novel severe acute respiratory syndrome coronavirus 2 (SARS-Cov-2) led to the beginning of a devastating pandemic that has disrupted the global economy and resulted in millions of deaths around the world [13]. Beyond the direct death resulting from COVID-19 infections, the pandemic has had a tremendous indirect impact due to overwhelmed healthcare systems and limited medical resources. Over two years have passed since the beginning of the pandemic, yet we do not fully understand the effects of public health mandates and policies on COVID-19 death and whether the vast difference in death counts between countries are attributable to regional intrinsic factors. In this study, we take a data-driven approach to understand this problem by developing a statistical modeling framework to identify key drivers of excess death and assess the effect of actionable factors like COVID-19 vaccination and policies. We show that intrinsic factors like age distribution and comorbidity risks that have been shown to be positively associated with COVID-19 mortality in prior studies are important in explaining the cross-country variation in excess mortality. We also show in accordance with past literature that certain intrinsic trust metrics are important when estimating excess mortality. Particularly, the trust the population has in its nation’s hospitals — potentially signifying a robust system more capable of handling the pandemic’s burden — is an important feature.

We further demonstrate the importance of actionable features including COVID-19 vaccination rate, policies, and trust. In particular, proportion of population with one COVID-19 vaccine dose and trust in COVID-19 advice from governments emerge at the top of the list for factors contributing to the predictive performance of our final model. Both of these variables show negative association with excess death. In addition, we detail the particularly deleterious effect that poor performance in these actionable variables can have on certain groups of countries. For instance, we show that low GDP countries experience a disproportionate contribution to estimated excess death if its citizens also do not trust their government’s pandemic-related advice. Similarly, we display that relatively obese countries experience a disproportionate contribution to estimated excess death if the country is relatively unvaccinated, and we quantify this effect. We also note that the two principle component features signifying the swiftness in the government’s response to the pandemic and the duration of such policy implementations did not appear near the top drivers of excess death. One explanation for this relatively low feature importance could be that there is a difference between policy implementation and policy compliance. This is perhaps too why the amount to which citizens trust their government’s pandemic-related advice appears as one of the most significant variables in explaining excess death.

While our study aims to be holistic, a number of limitations are present. For example, it is difficult to discern whether a government’s pandemic-related policy implementations were reactive or proactive in terms of excess death without including time in the analysis. Our future work for this study includes a temporal analysis that enables for a quantification of how different policies and intrinsic variables become significant or insignificant over time. One could also focus on a specific region of the world, which would lead to more complete data. Regardless, understanding the cross-country heterogeneity of excess death is key to gaining a comprehensive understanding of the true impact of the pandemic and can lead to actionable guidance for government and public health institutions in preventing future deaths. This study helps elucidate the factors contributing to excess mortality at the population level during the COVID-19 pandemic and can help guide governments in improving their response to pandemics in the future, ultimately saving human lives.

## 4 Methods

In this section, we give details of our overall modeling framework for our analysis and how we arrive at our “final model”. We build two models: one with only intrinsic features and one with both intrinsic and actionable features. We use and evaluate the predictions from these two models to assess the effects of actionable features such as vaccination rate and government policies during the COVID-19 pandemic.

### 4.1 Data collection and feature descriptions

First, we note that the time frame of our analysis is January 1, 2020 to November 1, 2021. This period is chosen for a number of reasons, including that the Omicron variant of the coronavirus SARS-CoV-2 was detected in early November 2021 [14], and different variants can contain varying patterns of spread and mortality. Additionally, this time frame still allows for the analysis of COVID-19 vaccination policies. For this study, we use the excess death estimates from WHO as the dependent variable. We obtain 34 different initial covariates for our analysis. Obesity percentage, age distribution, GDP per capita, population density, and hospital beds per 1000 people are a few of the “intrinsic” variables that were collected. Conversely, some of the collected features that are seen as at least moderately controllable over the course of the pandemic include number of days with comprehensive contact tracing, masking policies, workplace closures, days with available public testing, and official vaccination policies. The policy variables at the country-level were obtained from [15], while the socio-demographic, health, and government-spending habit information was obtained from World Bank Open Data. In addition to these data sources, we also include thorough survey information regarding trust of various entities among citizens for each country. We include these trust metrics since a number of studies report an association between trust and compliance to policy measures. For example, the study in [16] reports greater compliance for high-trust nations in the beginning of the pandemic, while [17] found an association between interpersonal trust and physical distancing adherence. Some of the trust metrics included in our analysis - such as overall confidence in the country’s hospitals and trust in the national government - are classified as intrinsic variables. Others - such as the degree to which citizens trust their government’s advice relating to COVID-19 measures - are classified as actionable. The survey data was obtained from [18]. COVID-19 related questions were typically asked in 2020, while other information to gauge citizen trust in various entities was obtained from 2018 survey data. For each survey, each country typically has approximately 1000 respondents. See Appendix A for the complete dataset description.

### 4.2. Final model construction for COVID-19 excess deaths

#### 4.2.1 Pre-processing

First, we note that some countries in our dataset did not have every covariate available. For countries that only have a few missing features, we impute these missing values via nuclear norm regularization. This method iteratively finds the soft-thresholded SVD and the algorithm in [19] is employed. After removing countries with many covariates missing and filtering for countries with over 5, 000, 000 people due to COVID-19 data quality concerns, we continue our analysis with 80 countries from around the world. Next, we note that interpretations of feature importance can be obfuscated when multi-collinearity is present, as it becomes challenging to rank variables in relative importance if a collection of such variables have similar effects. To avoid obscuring such interpretations and ensure drivers of excess death in the model can be reliably interpreted, we perform Principal Component Analysis (PCA) on the set of highly correlated features. The first principal component can be used in the model as a single variable representing the set of features as a linear combination. After creating various correlation plots, we decide to perform this method for variables falling into two groups: i) how quickly the country’s government responded to COVID-19 and ii) how long the country’s government held strict COVID-19 policies in place. An example of this dimension-reduction method for our dataset can be found in Figure 7. Critically, we note in Figure 7 that interpretations after this method are still clear. Since each feature contributes in the same direction along the first principal component, the newly created feature can be interpreted as “a greater, positive magnitude indicates a longer-lasting strictness in COVID-19 policy implementations”. We note too that when applying PCA to the second group of features with high multicollinearity - i.e. features relating to how quickly the government responded to the pandemic and put policies in place - all features also contribute in the same direction. This leads to the interpretation “a greater, positive magnitude indicates the government was swift in implementing stricter COVID-19 policies”.

**Fig. 7.**
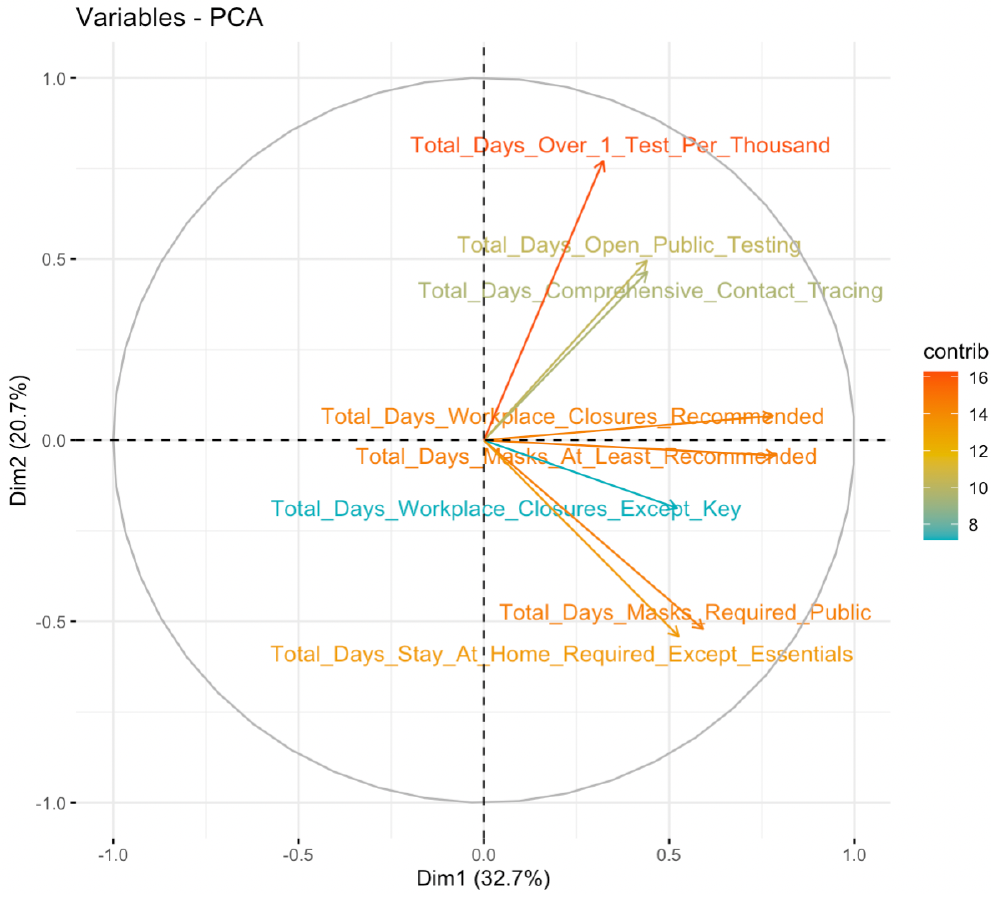
Bi-plot for one example of PCA in the employed dataset. All variables relate to how long the government’s COVID-19 policy was kept in place.

#### 4.2.2 Hyperparameter Tuning

After preprocessing the data, a number of models are employed using both the intrinsic and actionable features in the dataset. Specifically, Least Absolute Shrinkage and Selection Operator (LASSO) regression [20], Random Forest regression [21], and Gradient Boosting regression [22] models are created and analyzed. However, the limited sample size present in our dataset prevents the typical training and testing data split. Therefore, in order to estimate test accuracy for each model class and to pick hyperparameter combinations, we construct a repeated cross-validation scheme.

We collect 100 different CV predictions for each observation to obtain a more robust sense of predictions that could follow from slightly varying training sets. Figure 8 depicts the flow of the CV scheme.

**Fig. 8.**
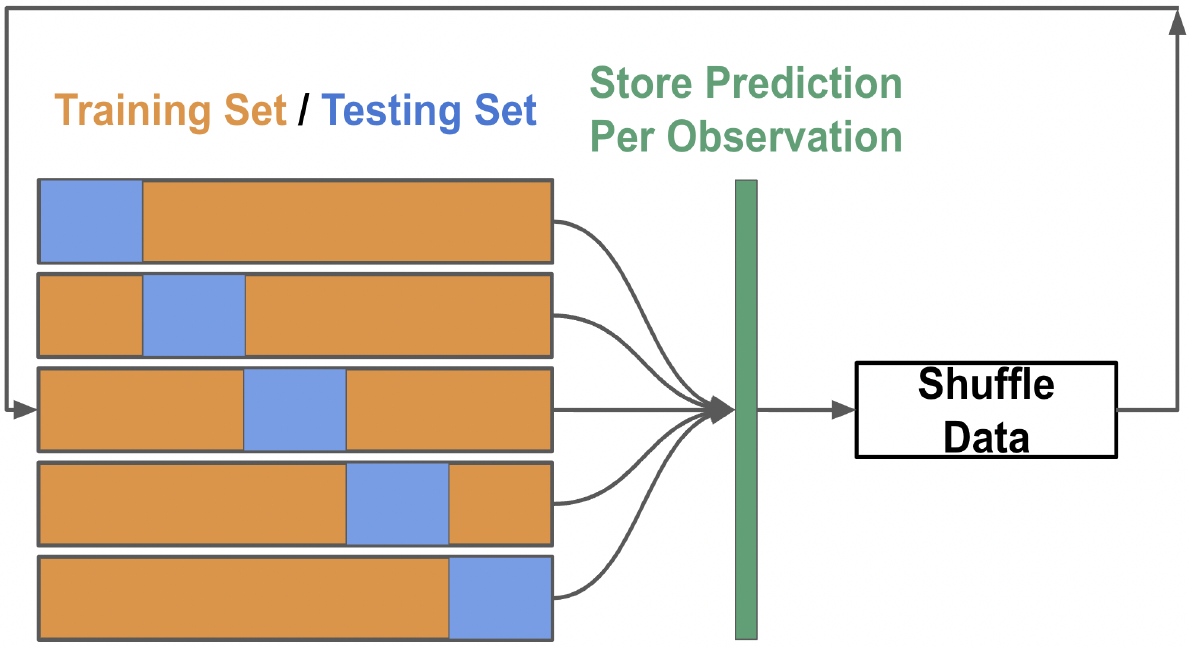
Schematic of the employed cross-validation algorithm. 100 predictions per observation were made for each hyperparameter combination

#### 4.2.3 Cross-Validation Results and Model Selection

Table 2 details the output from the 5-fold cross-validation scheme. We see that across the 100 repetitions, gradient boosting is the class of models that result in the lowest cross-validation (CV) error. The CV root mean squared error (RMSE) values in Table 1 involve all *N* * 100 values (resulting from 100 repetitions of 5-fold cross validation). Figure 9 details the distributions of the RMSE for each individual repetition of the repeated CV process. In Figure 9 we see that the gradient boosting model class with the specified hyperparameters consistently provides lower RMSE values for individual 5-fold CV trials. Due to this overall performance and consistency demonstrated through the repeated CV procedure, we choose to employ gradient boosting with the tuned hyperparameters as the “final model”.

**Fig. 9.**
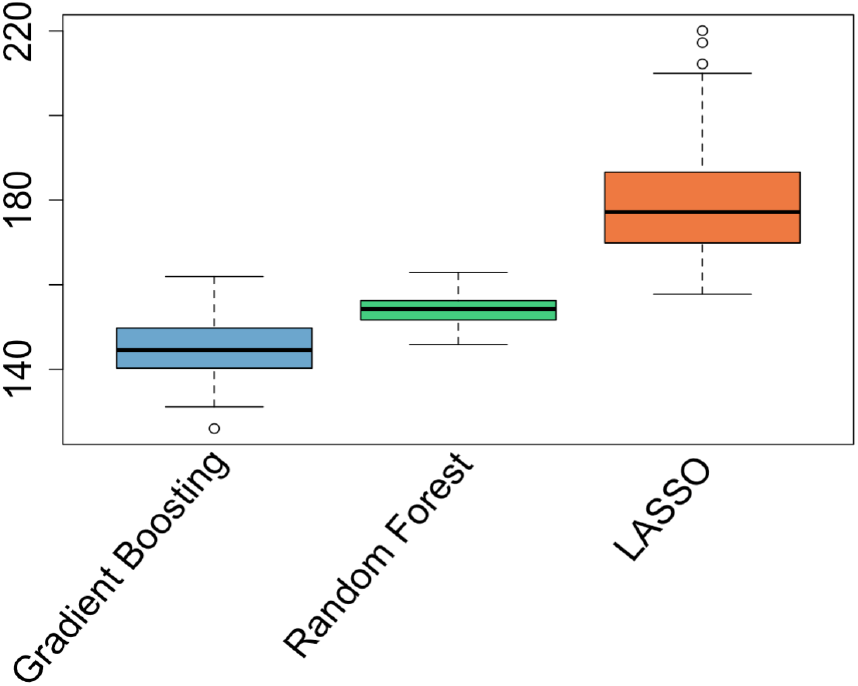
Distribution of CV RMSE. Each boxplot consists of the 100 different, individual CV RMSE values.

After seeing how the gradient boosting model class characterized by the hyperparameter values specified in Table 1 leads to lower CV error, the next step is to investigate how this type of model makes predictions. Therefore, we train a “final model” using all of the available data to get a more holistic sense of the model’s capturing of the inherent structure of the dataset. This “final model” leads to the insights discussed in section 2.

#### 4.2.4 Feature Importance and Partial Dependencies

The parameters and estimation process of the final model detailed in section 4.2.3 are dissected in numerous ways. Namely, feature importance and partial dependencies provide insights regarding the model. In our analysis, the importance of a feature is calculated via the increase in prediction error resulting from randomly permuting the values of the feature. The algorithm is similar to that in [21] but uses the entire dataset rather than out-of-bag observations [23]. Next, to define partial dependency, consider a sub-vector of input variables of interest denoted *χ_S_*, its complement with other inputs *χ_c_*, and an estimator 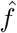. Partial dependency is defined as the marginal average [24]:

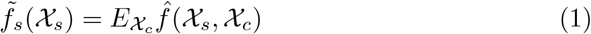

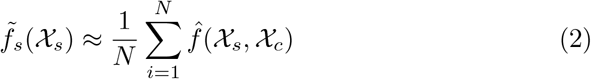

This accounts for the effect of input variables X_*s*_ after accounting for the average effects of the other variables *χ_c_*.

### 4.3 Bootstrap Hypothesis Testing

In order to evaluate the impact of the actionable features, we build two models following the same hyperparameter tuning procedure. One model is only fit on the set of intrinsic features while the other model is fit on the set of both the intrinsic and actionable features as shown in Figure 10. This dichotomy of models generates two sets of excess death predictions and RMSE. However, supervised learning algorithms can improve their predictive performance by increasing the number of features. In order to demonstrate that the set of actionable features are improving the model performance beyond the effect of simply adding more variables, we test the following hypothesis:

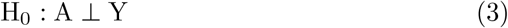

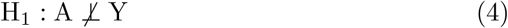

where A is the set of actionable features and Y is the response variable, excess death. To construct the null distribution, we perform the following bootstrapping procedure. We first fit a gradient boosting model with just the intrinsic features and compute the residuals. We bootstrap these residuals to generate 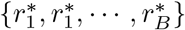. We add these residuals to the predicted excess death from the intrinsic-only model to get *y*^*^=*ŷ*_*I*_+*r*^*^. We fit another gradient boosting model with both intrinsic and actionable features and the corrupted response variable *y*^*^ and generate the residuals 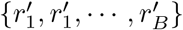. Finally we compute the difference of RMSE from the residuals *r*^*^ and *r*^′^ as our test statistic.

**Fig. 10.**
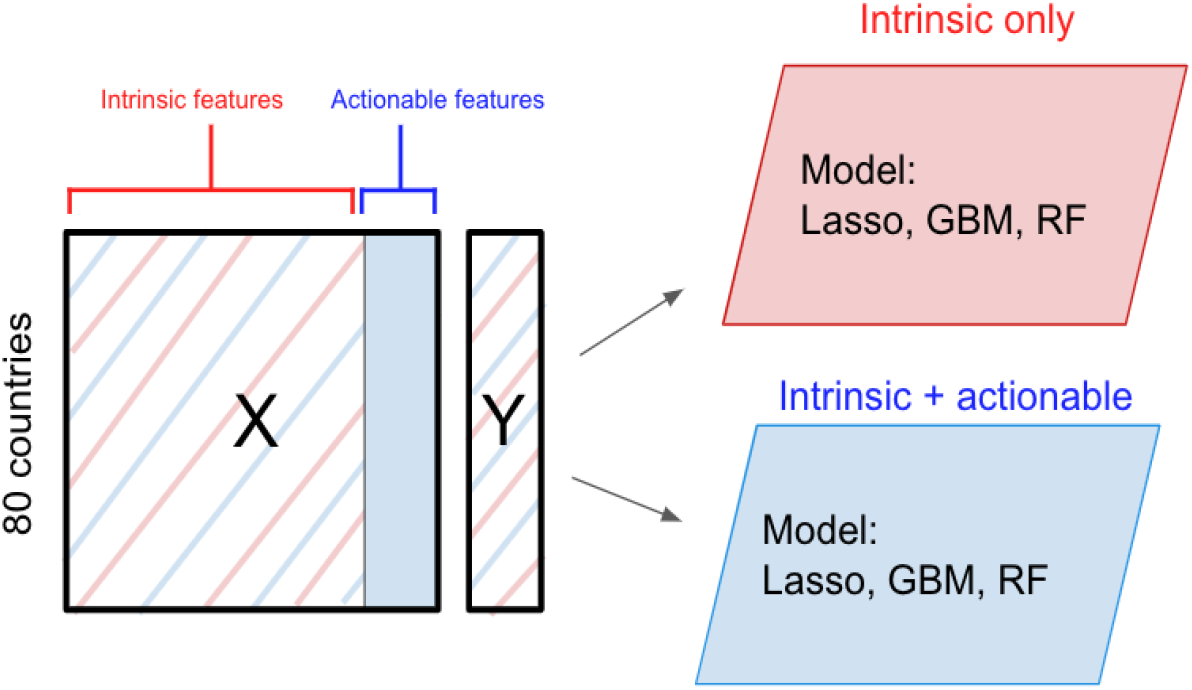
Diagram of the intrinsic vs. actionable feature modeling framework. Red represents the intrinsic features while blue represents the actionable features.

We repeat this procedure for 1000 iterations to generate 1000 bootstrapped RMSE differences. The p-value is given by,

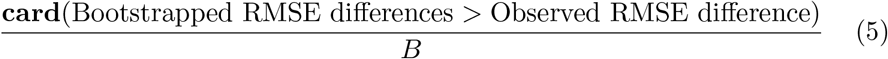

We attain a p-value of 0.0001 which suggests that the improvement in model performance through the inclusion of actionable features is statistically significant. The null distribution and observed RMSE difference is shown in Figure 11.

**Fig. 11.**
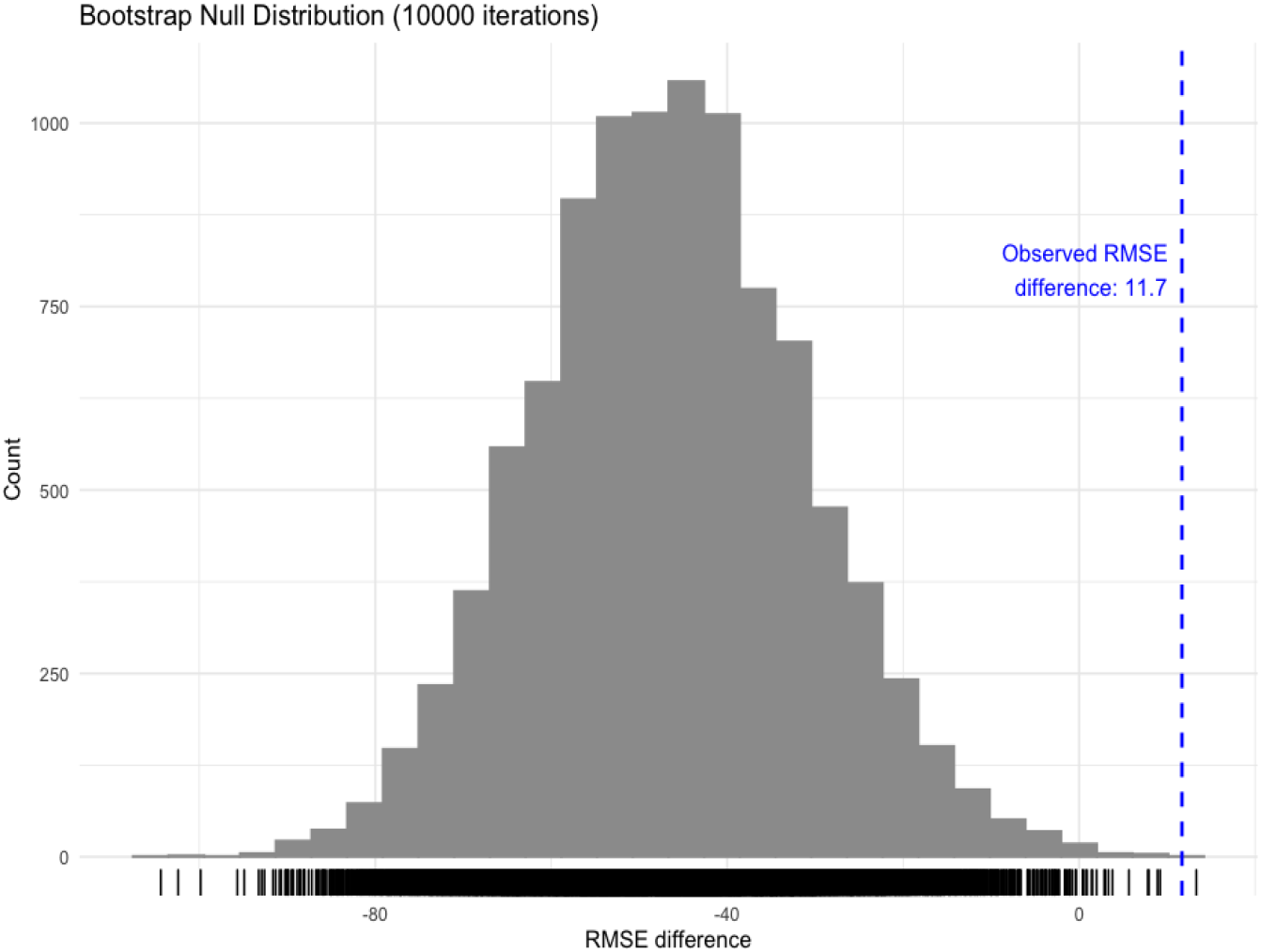
The gray histogram represents the null distribution constructed from 10,000 permuted RMSE differences and the red line represents the observed RMSE difference. This procedure results in a p-value of 0.0001.

### 4.4 Measuring the Effect of Actionable Features

The overall RMSE decrease after including the actionable features demonstrates the importance of these variables. To assess the effect of adding actionable features in the model for each country, we can measure how much the prediction changes between the model including and the model excluding the set of actionable features. Here, we compute the quantity for each country *i*:

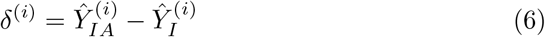

where *Ŷ*_*IA*_ are the excess death predictions from the model trained with both the intrinsic and actionable variables, and *Ŷ*_*I*_ are the excess death predictions from the model without the actionable variables.

We further construct 95% confidence intervals for delta estimates. We estimate the variance of the deltas by bootstrapping - i.e. sampling with replacement - the countries in the pre-processed data matrix and then computing the difference of excess death predictions from the two models *Ŷ*_*I*_ and *Ŷ*_*IA*_. We repeat this procedure for *B* iterations, generating *B* deltas 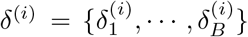 for each country *i*. We compute the sample standard deviation as:

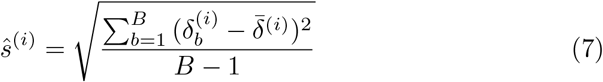

Using this estimate, we construct the confidence intervals for each country *i* as follows:

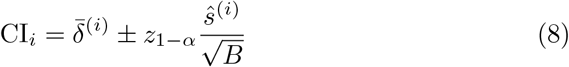

In practice due to sampling with replacement, some of the bootstrapped data matrix will have missing countries, so we bootstrap until each country has at least *B* number of *δ*.

## Data Availability

All data produced are available online at https://github.com/minwoosun/covid-mortality

## Appendix A Initial Dataset Description

**Table A1.**
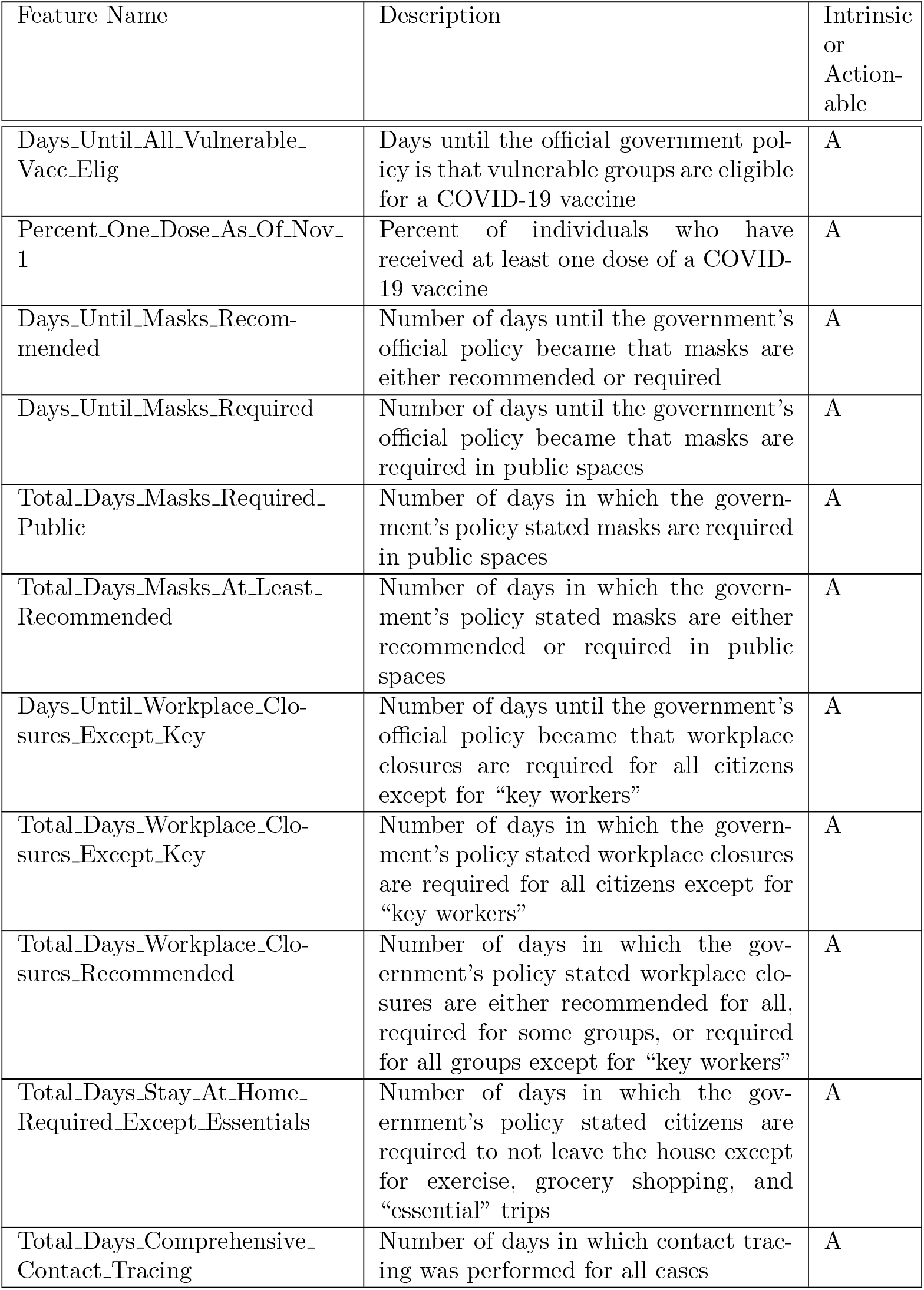
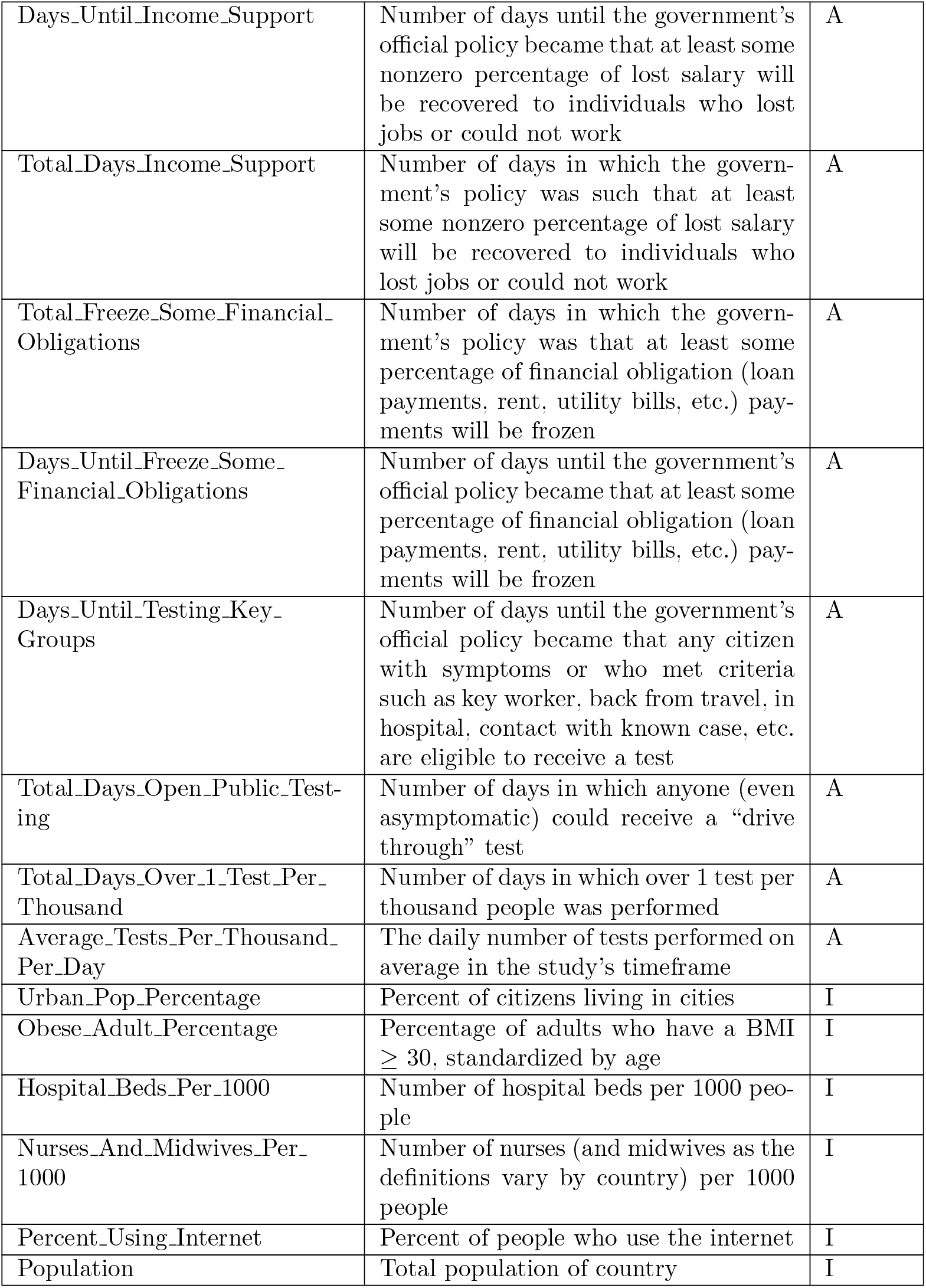
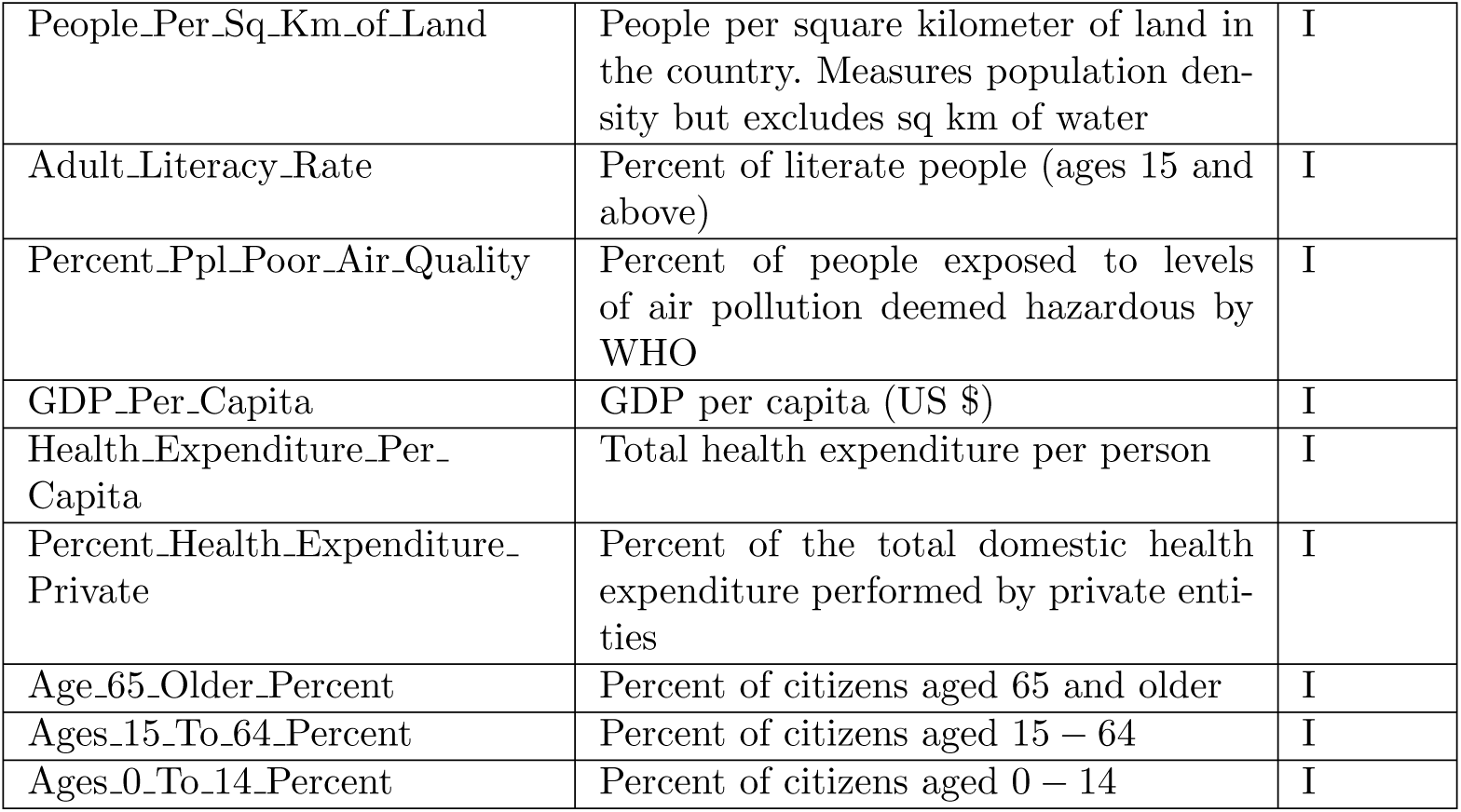
List of features for the initial dataset prior to any processing

