## Supplemental File 1 for "Do Public Health Efforts Matter? Explaining Cross-Country Heterogeneity in Excess Death During the COVID-19 Pandemic" (from another version of this preprint: version 1, 2022-11-22)

### Description of Features of Current Version of Country-Level Dataset

**Note:** We are confining this study from January 1st, 2020 to November 1st,2021 (approximate beginning of Omicron variant). Whether specified or not in this document, all time-related data has results only within this timeframe. This file includes all variables in the initial dataset before any filtering to reduce multicollinearity or for other reasons

Related to Response Variable

- cumulative_daily_covid_deaths_per100k: number of deaths attributed to COVID up until November 1, 2021.
- cumulative_daily_excess_deaths_per100k: number of estimated excess deaths from COVID up until November 1, 2021.
- cumulative_daily_excess_deaths_per100k_95lo: 95% confidence interval lower bound on the number of excess deaths within the specified timeframe.
- cumulative_daily_excess_deaths_per100k_95hi: 95% confidence interval upper bound on the number of excess deaths within the specified timeframe.

Vaccines and Cases

- Days_Until_All_Vulnerable_Vacc_Elig: Starting from January 1st, 2020, the number of days until the government's official policy became that key workers and clinically vulnerable groups and elderly groups are all eligible to receive a vaccine
  - Site: https://ourworldindata.org/covid-vaccination-policy
- Percent_One_Dose_As_Of_Nov_1: Percent of individuals who have received at least one dose of vaccine as of November 1st, 2021.
  - Site: https://ourworldindata.org/covid-vaccinations
- Total_Cases_Per_1000_People: Number of total positive recorded cases as of November 1st, 2021 per 1000 people living in country
  - Site: https://ourworldindata.org/covid-cases

Government Policies

- Days_Until_Masks_Recommended: Starting from January 1st, 2020, the number of days until the government's official policy became that masks are either recommended or required (i.e. at least recommended)
  - Site: https://ourworldindata.org/covid-face-coverings
- Days_Until_Masks_Required: Starting from January 1st, 2020, the number of days until the government's official policy became that masks are required in public spaces
  - Site: https://ourworldindata.org/covid-face-coverings
- Total_Days_Masks_Required_Public: Number of days starting from January 1st, 2020 until November 1st, 2021 in which the government's policy stated masks are required in public spaces

    - Site: https://ourworldindata.org/covid-face-coverings
- Total_Days_Masks_At_Least_Recommended: Number of days starting from January 1st, 2020 until November 1st, 2021 in which the government's policy stated masks are either recommended or required in public spaces
    - Site: https://ourworldindata.org/covid-face-coverings
- Days_Until_Workplace_Closures_Except_Key: Starting from January 1st, 2020, the number of days until the government's official policy became that workplace closures are required for all citizens except for "key workers"
    - Site: https://ourworldindata.org/covid-school-workplace-closures
- Total_Days_Workplace_Closures_Except_Key: Number of days starting from January 1st, 2020 until November 1st, 2021 in which the government's policy stated workplace closures are required for all citizens except for "key workers"
    - Site: https://ourworldindata.org/covid-school-workplace-closures
- Total_Days_Workplace_Closures_Recommended: Number of days starting from January 1st, 2020 until November 1st, 2021 in which the government's policy stated workplace closures are either recommended for all, required for some groups, or required for all groups except for "key workers" (i.e. at least recommended)
    - Site: https://ourworldindata.org/covid-school-workplace-closures
- Total_Days_Stay_At_Home_Required_Except_Essentials: Number of days starting from January 1st, 2020 until November 1st, 2021 in which the government's policy stated citizens are required to not leave the house except for exercise, grocery shopping, and "essential" trips
    - Site: https://ourworldindata.org/covid-stay-home-restrictions
- Total_Days_Comprehensive_Contact_Tracing: Number of days starting from January 1st, 2020 until November 1st, 2021 in which contact tracing was performed for all cases:
    - Site: https://ourworldindata.org/covid-testing-contact-tracing
- Avg_Gov_Stringency_Index: Average measure of strictness of government's policies from January 1st, 2020 to November 1st, 2021
    - Site: https://ourworldindata.org/covid-stringency-index
- Days_Until_Income_Support: Starting from January 1st, 2020, the number of days until the government's official policy became that at least some nonzero percentage of lost salary will be recovered to individuals who lost jobs or could not work:
    - Site: https://ourworldindata.org/covid-income-support-debt-relief
- Total_Days_Income_Support: Number of days starting from January 1st, 2020 until November 1st, 2021 in which the government's policy was such that at least some nonzero percentage of lost salary will be recovered to individuals who lost jobs or could not work:
    - Site: https://ourworldindata.org/covid-income-support-debt-relief
- Total_Freeze_Some_Financial_Obligations: Number of days starting from January 1st, 2020 until November 1st, 2021 in which the government's policy was that at least some percentage of financial obligation (loan payments, rent, utility bills, etc) payments will be frozen.
    - Site: https://ourworldindata.org/covid-income-support-debt-relief

- Days_Until_Freeze_Some_Financial_Obligations: Starting from January 1st, 2020, the number of days until the government's official policy became that at least some percentage of financial obligation (loan payments, rent, utility bills, etc) payments will be frozen.
    - Site: https://ourworldindata.org/covid-income-support-debt-relief

Tests

- Days_Until_Testing_Key_Groups: Starting from January 1st, 2020, the number of days until the government's official policy became that any citizen with symptoms or who met criteria such as key worker, back from travel, in hospital, contact with known case, etc. are eligible to receive a test:
    - Site: https://ourworldindata.org/covid-testing-contact-tracing
- Total_Days_Open_Public_Testing: Number of days starting from January 1st, 2020 until November 1st, 2021 in which any citizen (even asymptomatic) could receive a "drive through" test
    - Site: https://ourworldindata.org/covid-testing-contact-tracing
- Total_Days_Over_1_Test_Per_Thousand: Number of days starting from January 1st, 2020 until November 1st, 2021 in which over 1 test per thousand people was performed
    - Site: https://ourworldindata.org/coronavirus-testing
- Average_Tests_Per_Thousand_Per_Day: The daily number of tests performed on average from January 1st, 2020 to November 1st, 2021.
    - Derived from https://ourworldindata.org/coronavirus-testing. May not be exact as very crude extrapolation was used. (i.e. formula = (670-(days until first test))*(average tests per thousand per day in which we have data)/670). 670 is number of days in timeframe

Country-level Statistics

- Urban_Pop_Percentage: Percent of citizens living in cities
    - Site: https://data.worldbank.org/indicator/SP.URB.TOTL.IN.ZS
- Obese_Adult_Percentage: Percentage of adults who have a BMI >= 30, standardized by age
    - Site: https://ourworldindata.org/obesity
- Hospital_Beds_Per_1000: Number of hospital beds per 1000 people:
    - Site: https://data.worldbank.org/indicator/SH.MED.BEDS.ZS
- Nurses_And_Midwives_Per_1000: Number of nurses (and midwives as the definitions vary by country) per 1000 people:
    - Site: https://data.worldbank.org/indicator/SH.MED.NUMW.P3
- Percent_Using_Internet: Percent of people who use the internet:
    - Site: https://data.worldbank.org/indicator/IT.NET.USER.ZS
- Population: Total population of country:
    - Site: https://data.worldbank.org/indicator/SP.POP.TOTL

- People_Per_Sq_Km_of_Land: People per square kilometer of land in country. Measures population density but excludes sq km of water
    - Site: https://data.worldbank.org/indicator/EN.POP.DNST
- Adult_Literacy_Rate: Percent of literate people aged 15 and above
    - https://data.worldbank.org/indicator/SE.ADT.LITR.ZS
- Percent_Ppl_Poor_Air_Quality: Percent of people exposed to levels of air pollution deemed hazardous by WHO
    - Site: https://data.worldbank.org/indicator/EN.ATM.PM25.MC.ZS
- GDP_Per_Capita: GDP per capita (US $)
    - Site: https://data.worldbank.org/indicator/NY.GDP.PCAP.CD
- Health_Expenditure_Per_Capita: Total health expenditure per person
    - Site: https://data.worldbank.org/indicator/SH.XPD.CHEX.PC.CD
- Percent_Health_Expenditure_Private: Percent of the total domestic health expenditure performed by private entities
    - Site: https://data.worldbank.org/indicator/SH.XPD.PVTD.CH.ZS
- Age_65_Older_Percent: Percent of citizens aged 65 and older:
    - Site: https://data.worldbank.org/indicator/SP.POP.65UP.TO.ZS
- Ages_15_To_64_Percent: Percent of citizens aged 15-64
    - Site: https://data.worldbank.org/indicator/SP.POP.1564.TO.ZS
- Ages_0_To_14_Percent: Percent of citizens aged 0-14
    - Site: https://data.worldbank.org/indicator/SP.POP.0014.TO.ZS

Trust Variables

- Trust_Covid_Advice_Govt: Percent of people who base "some" or "a lot" of coronavirus decisions on advice from their national government
    - Site: https://wellcome.org/reports/wellcome-global-monitor-covid-19/2020 ("COVID-19 report: Crosstabs…")
- Trust_In_Neighborhood: Percent of people who have "some" or "a lot" of trust in the people in their neighborhood
    - Site: https://wellcome.org/reports/wellcome-global-monitor/2018 (Appendix D)
- Vaccines_Safe_50Plus: Percent of people aged 50+ who "somewhat agree" or "strongly agree" with the statement "Vaccines are safe."
    - Site: https://wellcome.org/reports/wellcome-global-monitor/2018 (Appendix D)
- Confidence_In_Hospitals: Percent of people who stated they have confidence in their nation's hospitals and health clinics
    - Site: https://wellcome.org/reports/wellcome-global-monitor/2018 (Appendix D)
- Trust_In_Govt: Percent of people who have "some" or "a lot" of trust in their national government
    - Site: https://wellcome.org/reports/wellcome-global-monitor/2018 (Appendix D)
- Trust_In_Journalists: Percent of people who have "some" or "a lot" of trust in journalists in their country
    - Site: https://wellcome.org/reports/wellcome-global-monitor/2018 (Appendix D)
